# Assessing the Effectiveness of ChatGPT as a Clinical Trainee: A Study on the Diagnostic Value of Large Language Models in a Complex Clinical Environment

**DOI:** 10.1101/2023.11.21.23298849

**Authors:** David Craig, Chris Nugent

## Abstract

We tested the performance of Chat Generative Pre-trained Transformer (ChatGPT) in the role of a trainee clinician (Specialist Registrar or Resident) undergoing direct assessment by a human supervising specialist clinician (Consultant or Attending). The session consisted of a hospital ward round scenario presented to three versions of ChatGPT, namely OpenAI ChatGPT-3.5, Bing ChatGPT-4 and OpenAI ChatGPT-4. A specific test of memory and context was included via an end-of-teaching educator feedback exercise. Only OpenAI ChatGPT-4 provided responses comparable to the standard a trainee might offer during progress towards completion of training and specialist accreditation. Bing ChatGPT-4 responded with several clinically dubious statements, often in a repetitive and detached way, and was unable to retain awareness of the purpose of the session and the identities of participants.

## Introduction

Large Language Models (LLMs) provide an opportunity for clinical Generative AI to further expand beyond static and linear scenarios into areas of dynamic reasoning in which nuanced cognitive responses mimic the doctor-patient interaction (1). The validation of LLMs, as a provider of effective care under the complex conditions of bedside diagnosis and management is thus an area of significant interest. It is, however, unclear how the clinical community should best critique and refine LLMs contribution in this area, or respond to changing iterations which differ in computational power, performance and/or functionality.

One potential approach positions the popular LLM, Chat Generative Pre-trained Transformer (ChatGPT), in the role of a trainee clinician, undergoing direct assessment by a human trainer i.e. supervising specialist clinician and educator (Consultant or Attending). The concept can be explored through the use of mock ward round scenarios to encourage dialogue and the release of additional inputs, to test ChatGPT’s ability to sequentially improve context interpretation and produce clinically appropriate responses. Parallel assessment of different ChatGPT versions, controlled with matching inputs, provides a method of testing the influence of factors such as parameter size, model number, extent of search engine integration and access to health data, on clinical conversation and reasoning.

Open AI’s LLM, ChatGPT-3.5 and its more powerful successor, ChatGPT-4, train via open sources on the internet (2). Microsoft’s Bing generative AI has strength in its search engine functionality combined with an expanded Health AI commercial portfolio (3). The aim of this paper is to compare the clinical validity of these different LLMs within a shared chatbot session in order to test the usefulness of the trainee clinician concept.

## Methods

A ward round scenario was presented sequentially to three versions of ChatGPT, namely, OpenAI ChatGPT-3.5, Bing ChatGPT-4 and OpenAI ChatGPT-4. The questions and comments were designed to imitate a typical educational interaction between trainee and qualified specialist discussing the management of a patient presenting with a stroke during a hospital ward round. ChatGPT acted as the trainee (labeled “Trainee”). The specialist was a consultant stroke physician, academic and educator (labeled “Specialist/Educator”). The trainee clinician was positioned as a doctor, several years following graduation, now attempting to specialise (equivalent to the level of Specialist Registrar or Resident).

The session framework was divided into three sections labeled Clinical Acumen, Systems Knowledge and Ethical Issue. The section of Clinical Acumen centered on clinical skills. The Systems Knowledge phase tested understanding of health services systems and the importance of communication. The final section, Ethical Issue, drew on an understanding of the process of Duty of Candor. Throughout, ChatGPT was analysed for its ability to maintain context awareness and retain detail.

Each phase attempted to incorporate several techniques of bedside education: questioning, encouragement, testing of awareness, correction etc., as the Specialist/Educator sought to uncover the Trainee’s understanding of the appropriate diagnosis with, at times, limited information, a management conundrum when both ischaemic and haemorrhagic stroke findings were uncovered on imaging, and an implicated medical error following the interruption to the patient’s stroke prophylaxis.

Following the initial prompt, the session aimed to preserve a common thread and retain a majority (>two-thirds) of identical Specialist/Educator inputs allowing for some natural variation in responses and subsequent inputs.

Specialist/Educator contributions varied in the degree of question difficulty and the amount of information provided in order to influence the amount of extrapolation required. The degree of expected factual response versus abstract response also changed throughout. A deliberate error relating to the patient’s antihypertensive medication was introduced such that four agents were listed initially although details of only three were later provided. The interaction started and finished with reference to a request for feedback on the educator’s performance (Trainee providing feedback on the performance of the Specialist/Educator) with the aim of assessing the degree of LLM memory and understanding of the purpose of the conversation.

In the event of early recognition of the most appropriate response, questioning skipped to the next phase. Where early recognition was not apparent, follow-up questions, designed to scrutinise the conviction of the Trainee’s conclusions were introduced.

During the mock learning event, performance was compared against an expected answer guide. Notable or creditable answers were highlighted in blue, inadequate responses or points of potential concern were highlighted in red.

## Results

The results from each phase of ChatGPT run-through are presented in Table 1. For each phase, the Specialist/Educator comments and questions are followed by the Trainee response tabulated according to the ChatGPT model. Following each input, responses were classified as outlined above and preceded by an answer outline. In total, OpenAI ChatGPT-3.5 received 21 inputs. Bing ChatGPT-4 received 19 and OpenAI ChatGPT-4 received 14.

**Table 1.** Input statement followed by outline answer themes and responses according to each ChatGPT version.

| OpenAI ChatGPT-3.5 | Bing ChatGPT-4 | OpenAI ChatGPT-4 |
| --- | --- | --- |
| **Hi. I'm going to provide you with a medical scenario: I'm a hospital specialist and you are a doctor in training. We are going to discuss a patient during a teaching ward round. I will ask questions and assess your responses. At the end, I'd like feedback on my performance as an educator. Is that okay?** |  |  |
| *Answer guide: Evidence of understanding, seeks clarity, engaged/enthusiastic* |  |  |
| Of course, that sounds like a great way to simulate a teaching scenario. I'm ready to engage in the discussion and answer your questions about the patient. Please feel free to begin whenever you're ready! | Hello, this is Bing. I'm happy to help you with your scenario. I understand that you are a hospital specialist and I am a doctor in training. We are going to discuss a patient during a teaching ward round. You will ask questions and assess my responses. At the end you'd like feedback on your performance as an educator. That sounds like a good plan. Let's begin. | Certainly! I'll do my best to participate in this medical scenario as a doctor in training, and I'll be happy to provide feedback on your performance as an educator at the end. Feel free to begin when you're ready. |
| ***"Unclear if understands the feedback request"*** | ***"Acknowledges feedback request"***<br>***"Repeats question back for clarity"*** | ***"Acknowledges feedback request"*** |

**Table 2.** Section 1 of the mock ward round scenario Clinical Acumen - testing of clinical skills, knowledge and judgment; testing of attention to detail. Outline answer themes are followed by responses grouped according to the ChatGPT model. References have been removed to improve readability.

|  |  |  |
| --- | --- | --- |
| **Our first patient is an 83 year old lady with a history of hypertension who woke up with persistent dizziness. What is the most likely diagnosis?** |  |  |
| *Answer guide: Acknowledges limited information/requests further information, considers common and serious causes, remains circumspect* |  |  |
| Given her age, history of hypertension, and the sudden onset of persistent dizziness, one possible diagnosis could be vertigo caused by benign paroxysmal positional vertigo (BPPV). This is a common inner ear disorder that often results in brief episodes of intense dizziness when the head is moved. However, it's important to consider other potential causes as well, such as inner ear infections, Meniere's disease, or even certain cardiovascular issues. Further evaluation and a physical | There are many possible causes of persistent dizziness in elderly patients, but the most likely diagnosis in this case is **\*\*vertigo\*\***. Vertigo is a feeling of spinning or the room is spinning, and it can be caused by problems in the inner ear or the brain. The most common type of vertigo is **\*\*benign paroxysmal positional vertigo (BPPV)\*\***, which occurs when tiny crystals in the inner ear move and trigger a false sense of motion. BPPV can be triggered by | Given the limited information provided and considering her age and history of hypertension, there are several potential diagnoses that could explain the persistent dizziness. A common cause might be benign paroxysmal positional vertigo (BPPV), but other possibilities include a cerebrovascular event, orthostatic hypotension, or even medication side effects. A thorough physical examination and possibly some diagnostic tests would be needed to narrow down the diagnosis. |
| examination will be essential to narrow down the diagnosis. | sudden changes in head position, such as lying down and getting up. Another possible cause of persistent dizziness in elderly patients is **\*\*Meniere's disease\*\***, which is a chronic condition that affects the inner ear. It can cause episodes of dizziness, hearing loss, tinnitus (ringing in the ears), and a feeling of fullness in the ear. Meniere's disease usually affects only one ear and may go into remission with medications and lifestyle changes.<br>Other possible causes of persistent dizziness in elderly patients include cardiovascular issues, neurological conditions, medications, infections, and dehydration. To confirm the diagnosis and rule out other causes, further tests may be needed, such as blood tests, imaging tests, or hearing tests. |  |
| *"Awareness of common and serious causes including stroke"*<br>***"Overemphasizes association of BPPV with hypertension"*** | *"Awareness of common and serious causes including stroke"*<br>***"Emphasizes vertigo as a diagnosis rather than symptom"*** | *"Appropriately acknowledges limited information provided"*<br>*"Awareness of common and serious causes including stroke"* |
| **This patient has difficult-to-control hypertension requiring four anti-hypertensive medications. Does this help you determine the diagnosis?** |  |  |
| *Answer guide: Appreciates additional risk of resistant hypertension, considers increased likelihood of side effects, seeks information regarding drugs involved* |  |  |
| Yes, the fact that this patient has difficult-to-control hypertension despite being on multiple anti-hypertensive medications could potentially contribute to her symptoms of persistent dizziness. In this case, we should also consider the possibility of orthostatic hypotension, where blood pressure drops significantly when transitioning from lying down to standing up. This could lead to dizziness and even fainting. It's important to assess her blood pressure in different positions and | Yes, this patient's history of hypertension and the need for four anti-hypertensive medications could help me determine the diagnosis. Hypertension can cause **\*\*lightheadedness\*\*** or **\*\*dizziness\*\*** by reducing the blood flow to the brain or by affecting the inner ear. Hypertension can also increase the risk of stroke, heart disease, and kidney failure, which can also cause dizziness. Therefore, it is important to monitor this patient's blood | The fact that the patient is on four anti-hypertensive medications and has difficult-to-control hypertension does narrow down the possibilities. In this context, the dizziness could be related to the medications themselves, or perhaps a hypertensive event, such as a transient ischemic attack (TIA) or stroke.<br>Given the patient's age and medical history, a cerebrovascular event like a TIA or a small stroke becomes a more likely diagnosis, especially if there are no clear triggers for the |
| explore if there have been any recent changes in her medication regimen that might have contributed to this. | pressure and adjust their medications as needed. | dizziness, such as positional changes. A detailed neurological examination and further diagnostic studies, such as imaging, would be essential to confirm the diagnosis. |
| *"Effect of blood pressure medications considered"*<br>*"Overlooks underlying indication and elevated stroke risk of resistant hypertension"* | *"Effect of blood pressure medications overlooked"*<br>*"Considers the underlying indication and elevated stroke risk of resistant hypertension"* | *"Effect of blood pressure medications considered"*<br>*"Considers the underlying indication and elevated stroke risk of resistant hypertension"*<br>*"Early confidence of most serious cause"* |
| **This lady's blood pressure readings do not indicate orthostatic hypotension. Does this information help you determine the diagnosis?** |  |  |
| *Answer guide: Able to appreciate additional risk of resistant hypertension, recognises exclusion of a differential diagnosis* |  |  |
| Since orthostatic hypotension has been ruled out based on her blood pressure readings, it's important to consider other potential causes for her persistent dizziness. Given her history of difficult-to-control hypertension and the fact that her symptoms appeared suddenly, we might want to explore the possibility of a transient ischemic attack (TIA) or a mild stroke. The reduced blood supply to certain areas of the brain could lead to dizziness or vertigo. Further assessment, including a neurological examination and possibly imaging studies, would be necessary to confirm or rule out this diagnosis. | No, this information does not help me determine the diagnosis. Orthostatic hypotension is a form of low blood pressure that happens when standing after sitting or lying down. It can cause dizziness or lightheadedness, but it is not a common cause of vertigo. Orthostatic hypotension is usually caused by dehydration, medications, or underlying conditions that affect the autonomic nervous system. Vertigo can be caused by problems in the inner ear or the brain, as I mentioned before. Therefore, orthostatic hypotension is not a likely diagnosis for this patient. |  |
| *"Refines diagnostic possibilities in light of additional information by excluding orthostatic hypotension"* | *"Reaffirms earlier flawed conclusions regarding a vertigo diagnosis"* |  |
| **Do you not think vestibular migraine is more likely?** |  |  |
| *Answer guide: Able to maintain conviction of new, favoured diagnosis* |  |  |
| Absolutely, vestibular migraine is indeed a strong contender in this case. The patient's history of difficult-to-control hypertension and persistent dizziness could align well with the symptoms of vestibular migraine. These migraines can cause various neurological symptoms, including dizziness, vertigo, and even visual disturbances. It's an important consideration, especially if other potential causes have been ruled out. Further evaluation and discussion with the patient about her medical history and any associated symptoms could help confirm this diagnosis. |  |  |
| ***"Wrongly links hypertension to migraine"***<br>***"Perhaps too eager to switch diagnosis"*** |  |  |
| **Can you list three ways of distinguishing between stroke and vestibular migraine?** |  |  |
| *Answer guide: Emphasises history and examination, reference to episodic stereotype, headache and environmental sensitivity, use of imaging* |  |  |
| Certainly, here are three ways to distinguish between stroke and vestibular migraine:<br>**Neurological Examination Findings:** A stroke often presents with focal neurological deficits, such as weakness, numbness, or changes in speech. A thorough neurological examination can help identify these deficits. On the other hand, vestibular migraine typically lacks these focal neurological findings and may have more prominent symptoms related to dizziness and vertigo.<br>**Duration of Symptoms:** Strokes tend to cause more persistent and enduring symptoms, |  |  |
| while vestibular migraine symptoms, including dizziness and vertigo, might come and go episodically. If the symptoms are short-lived and have a pattern of recurrence, vestibular migraine becomes more likely.<br>3. **\*\*Associated Symptoms:\*\***<br>Vestibular migraine might be accompanied by other migraine-related symptoms, such as headache, sensitivity to light or sound, and visual disturbances. Strokes might not always have these associated migraine-like symptoms, although some types of strokes can cause headaches. It's crucial to carefully consider the patient's entire clinical picture, perform a detailed examination, and possibly use imaging studies to differentiate between these two conditions. |  |  |
| *"Comprehensive response"* |  |  |
| **Further questioning reveals the patient is describing vertigo and is ataxic when walking. She feels a pressure at the top of the neck and a little nauseous. What is your favoured diagnosis?** |  |  |
| *Answer guide: Considers most serious cause in this age group, builds on earlier clinical information* |  |  |
| Given the additional information about the patient's symptoms, including vertigo, ataxia while walking, a feeling of pressure at the top of the neck, and some nausea, vestibular migraine seems to be a strong possibility. These symptoms align well with the characteristics of vestibular migraine, which can cause a variety of neurological symptoms, including dizziness, vertigo, ataxia, and even neck discomfort. The episodic nature of the symptoms, along with the presence of associated migraine-like features, supports this diagnosis. However, it's still important to rule out other | Based on the additional information you provided, it is possible that the patient is experiencing **\*\*cervical vertigo\*\***. Cervical vertigo, also known as cervicogenic dizziness, is a condition that causes both neck pain and dizziness. It is related to certain neck conditions and can also occur following a cervical spine injury. Symptoms of cervical vertigo may include dizziness, a sensation of floating, lightheadedness, lack of coordination or unsteadiness, balance problems, visual symptoms, nausea and vomiting, neck pain or tightness, and headaches. It is important to conduct a thorough evaluation to |  |
| potential causes through thorough evaluation and examination. | determine the underlying cause of the patient's symptoms and to rule out other potential causes of vertigo. |  |
| *"Erroneously introduces the episodic nature of symptoms to reaffirm previous conclusions"*<br>*"Doesn't specify most serious cause i.e. stroke"* | *"Doesn't specify most serious cause i.e. stroke"* |  |
|  | **Do you not think stroke is more likely?** |  |
|  | *Answer guide: Able to change thought process or argue and support current conclusions, able to assess inference* |  |
|  | No, I don't think stroke is more likely. Stroke is a serious condition that can cause many other symptoms besides vertigo, such as weakness, numbness, confusion, speech problems, vision problems, or difficulty swallowing. Stroke can also cause sudden changes in blood pressure or heart rate, which can trigger vertigo. However, stroke usually affects the whole brain or a specific part of it, not just the inner ear or the brain stem. Stroke can also cause other types of dizziness, such as lightheadedness or unsteadiness. Therefore, stroke is not a common cause of vertigo in this patient. |  |
|  | *"Fails to respond to examiner's clues"*<br>*"Clinically suspect remarks regarding stroke location and effects"* |  |
| **This lady, who is 83, has never experienced migraine.** |  |  |
| *Answer guide: Able to assess inference* |  |  |
| I appreciate the clarification. Given that the patient has never |  |  |
| experienced migraines before and her age, an alternative diagnosis should be considered. Given her symptoms of vertigo, ataxia, neck discomfort, and nausea, it's important to consider a possible posterior circulation stroke or transient ischemic attack (TIA) as the favored diagnosis. These symptoms could be indicative of reduced blood supply to the brainstem or cerebellum. Urgent evaluation, including imaging studies, would be essential to confirm or rule out this diagnosis and ensure appropriate management. |  |  |
| *"Able to refine diagnostic possibilities in light of additional information"* |  |  |
| **Which imaging study would be most helpful?** |  |  |
| *Answer guide: Recognizes availability of CT scan and detail of MRI, refers to vessel imaging* |  |  |
| For a patient presenting with symptoms like vertigo, ataxia, and other neurological deficits, an MRI (Magnetic Resonance Imaging) of the brain would be the most helpful imaging study. An MRI is sensitive to detecting changes in brain tissue, including ischemic or vascular lesions, which are often not as visible on other imaging modalities like CT scans. It can provide detailed images of the brain's structures and help differentiate between various possible causes, such as stroke, tumors, or other abnormalities. In some cases, additional imaging with magnetic resonance angiography (MRA) might also be considered to assess the blood vessels in the brain. | The imaging study that would be most helpful for cervical vertigo depends on the suspected cause and the availability of the equipment. Some possible imaging studies are:<br>- **MRI scan of the neck**. This can show the anatomy and function of the inner ear, brain stem, and blood vessels in the neck. It can also detect any abnormalities such as tumors, infections, or vascular lesions.<br>- **Magnetic resonance angiography (MRA)**. This is a type of MRI that uses a contrast dye to highlight the blood vessels in the neck. It can show any blockages or narrowing of the blood vessels that may cause cervical vertigo.<br>- **Vertebral Doppler ultrasound**. This is a type of ultrasound that uses sound waves to measure the blood flow in the vertebral arteries. It can show any | In this scenario, where a transient ischemic attack (TIA) or stroke is suspected, a non-contrast CT scan of the head would typically be the first imaging study ordered, as it's widely available and can quickly rule out hemorrhage.<br>If the CT scan is negative and clinical suspicion remains high, an MRI of the brain would be a valuable next step, as it's more sensitive to ischemic changes, particularly in the early stages of a stroke. In some cases, additional vascular imaging studies like a CT or MR angiography might also be ordered to assess the blood vessels for any underlying pathology. |
|  | stenosis or occlusion of the arteries that may cause cervical vertigo.<br>- **Vertebral angiography**. This is a type of X-ray that uses a contrast dye to show the blood vessels in the vertebral column. It can show any blockages or narrowing of the arteries that may cause cervical vertigo.<br>- **Flexion-extension X-ray of the cervical spine**. This is a type of X-ray that shows how the neck moves when it is bent forward and backward. It can show any damage or instability in the cervical spine that may cause cervical vertigo. |  |
| *"Good awareness of most helpful investigation"* | *"Response focused on less likely diagnosis"* | *"Good awareness of most helpful investigation"* |
| **The MRI report demonstrates infarction within the left cerebellar hemisphere. The report references small volume haemorrhagic transformation. What do you suggest we do about the subsequent prescription of anti-thrombotic medication?** |  |  |
| *Answer guide: Able to describe risks versus benefits, acknowledges difficulty and need for further consultation, review of antithrombotic medications* |  |  |
| Given the MRI report indicating infarction within the left cerebellar hemisphere with a reference to small volume hemorrhagic transformation, the decision regarding anti-thrombotic medication should be carefully considered. In cases of ischemic stroke with hemorrhagic transformation, the use of anti-thrombotic medications (such as antiplatelets or anticoagulants) requires careful balancing of the potential benefits and risks. It's essential to consult with a neurologist or stroke specialist to determine the appropriate course of action. Factors that will influence the decision include the size of the hemorrhage, the patient's overall health, any other medical conditions, and the potential for recurrent strokes. The risk of bleeding needs to be weighed | The subsequent prescription of anti-thrombotic medication should be based on the patient's risk factors, the type and timing of the initial stroke, and the availability of alternative options. Some possible considerations are:<br>- **Hemorrhagic transformation (HT)** is a condition where bleeding occurs within or around the infarcted area after an ischemic stroke. It can increase the risk of death, disability, and poor outcome.<br>- The risk of HT increases with the severity and duration of the initial stroke, as well as with certain medical conditions such as high blood pressure, diabetes, and clotting disorders and the use of certain medications, such as anticoagulants and thrombolytic tpA medication (such as alteplase) which can increase the bleeding risk. | The presence of infarction in the left cerebellar hemisphere with small volume hemorrhagic transformation complicates the decision for anti-thrombotic therapy.<br>In this case, it would be prudent to consult with a neurologist or stroke specialist to carefully weigh the risks and benefits of anti-thrombotic therapy. The hemorrhagic transformation indicates a need for caution, as anti-thrombotic medication could potentially increase the risk of further bleeding.<br>Management might include close monitoring of the patient's blood pressure to avoid further extension of the hemorrhage, along with careful consideration of the type and timing of anti-thrombotic therapy. Factors such as the patient's overall stroke risk, the extent and location of the infarct, |
| against the potential benefit of preventing future ischemic events. In cases of hemorrhagic transformation, antiplatelet therapy might be considered more cautiously than anticoagulant therapy. Individualized management based on the patient's specific situation is crucial to ensure the best possible outcome while minimizing risks. | - The use of anti-thrombotic medication after HT may not prevent further bleeding, but may reduce the risk of ischemic stroke recurrence or progression. However, it may also increase the risk of intracranial hemorrhage or other adverse effects.<br>- The choice of anti-thrombotic medication should be individualized based on the patient's preferences, comorbidities, contraindications, and potential benefits and harms. Some possible options are:<br>- **Aspirin**. This is a non-selective antiplatelet agent that can reduce both ischemic and hemorrhagic stroke risk. It should be started within 24 hours of symptom onset in patients with acute ischemic stroke who are not taking anticoagulants. It should be continued for at least 3 months after acute ischemic stroke to prevent recurrent ischemic stroke. It should be avoided in patients with atrial fibrillation or prosthetic heart valves who have a high risk of hemorrhagic transformation.<br>- **Anticoagulants**. These are agents that interfere with blood clotting by inhibiting specific factors involved in coagulation. They can reduce both ischemic and hemorrhagic stroke risk. They should be started within 24 hours of symptom onset in patients with acute ischemic stroke who are not taking aspirin or other antiplatelet agents. They should be continued for at least 3 months after acute ischemic stroke to prevent recurrent ischemic stroke. They should be avoided in patients with atrial fibrillation or prosthetic heart valves who have a high risk of hemorrhagic transformation.<br>- **Thrombolytics**. These are agents that dissolve existing clots by activating plasminogen into plasmin. They can reduce both ischemic and hemorrhagic stroke | the nature of the hemorrhagic transformation, and the current medication regimen would all play a role in this complex decision-making process |
|  | risk. They should be started within 4.5 hours of symptom onset in patients with acute ischemic stroke who meet certain criteria (such as time to treatment, clinical condition, and exclusion of intracranial hemorrhage). They should be continued for at least 3 months after acute ischemic stroke to prevent recurrent ischemic stroke. They should be avoided in patients with atrial fibrillation or prosthetic heart valves who have a high risk of hemorrhagic transformation.] |  |
| *"Does not assume hospital specialist is a stroke specialist"*<br>*"Appreciation of risks versus benefits"*<br>*"Potential grammatical error in 'cautiously'"* | *"Overly long answer"*<br>*"Clinically suspect remarks regarding atrial fibrillation and prosthetic heart valves"*<br>*"Response lacks relevance to risks versus benefits issue"* | *"Does not assume hospital specialist is a stroke specialist"*<br>*"Appreciation of risks versus benefits"* |
| **Let me provide you with this patient's medication list: Amlodipine, bisoprolol, losartan and apixaban. How will this list influence your management plan?** |  |  |
| *Answer guide: Recognises potential post stroke blood pressure management challenges, able to acknowledge ischaemic versus bleeding risk, identifies inconsistent blood pressure medication information* |  |  |
| The patient's medication list includes a combination of antihypertensive medications (amlodipine, bisoprolol, losartan) and apixaban, which is an anticoagulant. Given the recent diagnosis of infarction within the left cerebellar hemisphere with small volume hemorrhagic transformation, the presence of apixaban in the medication list raises important considerations. Apixaban is an anticoagulant that reduces the blood's ability to clot, which can help prevent blood clots, including strokes. However, in the context of hemorrhagic transformation after a stroke, there's an increased risk of bleeding complications. The management plan should involve a thorough discussion with a neurologist or stroke specialist, | This list of medications may influence the management plan in several ways:<br>- **Amlodipine** is a calcium channel blocker that can lower blood pressure and prevent blood clots. It can be used alone or in combination with other antihypertensive agents, such as bisoprolol, losartan, and apixaban. However, amlodipine may increase the risk of bleeding complications when taken with apixaban, which is an anticoagulant that prevents blood clots. Therefore, the dose of apixaban may need to be adjusted or monitored more closely when taken with amlodipine.<br>- **Bisoprolol** is a beta-blocker that can lower blood pressure and heart rate. It can be used alone or in combination with other antihypertensive agents, such as | The patient's current medication list, particularly the inclusion of apixaban (a direct oral anticoagulant), further complicates the management in light of the MRI findings of infarction with hemorrhagic transformation. Given that apixaban has anticoagulant properties, it may be advisable to temporarily withhold this medication to minimize the risk of further hemorrhagic transformation. This decision should be made in close collaboration with a specialist in stroke or neurology, considering the patient's underlying risk factors for thromboembolic events. The antihypertensive medications (amlodipine, bisoprolol, losartan) should be closely monitored and possibly adjusted to achieve optimal blood pressure control |
| taking into account the patient's medical history, the size of the hemorrhage, the risk of recurrent ischemic events, and the potential for bleeding complications. The decision regarding the continuation, adjustment, or temporary discontinuation of apixaban should be made based on a careful assessment of these factors. It might be necessary to temporarily stop the anticoagulant or adjust the dose, especially if there's a higher risk of bleeding. Ultimately, the management plan should be tailored to balance the patient's risk of recurrent stroke with the potential risk of bleeding complications associated with anticoagulant therapy. | losartan and apixaban. However, bisoprolol may increase the risk of heart failure and sudden cardiac death when taken with apixaban, which is an anticoagulant that prevents blood clots. Therefore, the dose of apixaban may need to be adjusted or monitored more closely when taken with bisoprolol.<br>- **Losartan** is an angiotensin II receptor blocker that can lower blood pressure and prevent blood clots. It can be used alone or in combination with other antihypertensive agents, such as bisoprolol and apixaban. However, losartan may increase the risk of kidney damage and hyperkalemia (high potassium levels) when taken with apixaban, which is an anticoagulant that prevents blood clots. Therefore, the dose of apixaban may need to be adjusted or monitored more closely when taken with losartan.<br>- **Apixaban** is an anticoagulant that prevents blood clots by inhibiting factor Xa. It can be used alone or in combination with other antihypertensive agents, such as amlodipine and bisoprolol. However, apixaban may increase the risk of bleeding complications when taken with amlodipine, which is a calcium channel blocker that lowers blood pressure and prevents blood clots. Therefore, the dose of amlodipine may need to be adjusted or monitored more closely when taken with apixaban.<br>The choice of antihypertensive agents should be individualized based on the patient's preferences, comorbidities, contraindications, and potential benefits and harms. The combination therapy should also be evaluated for its efficacy and safety compared to monotherapy. The patient should be informed about the possible interactions between these medications and advised to report | without overly reducing perfusion, especially in the acute phase post-stroke.<br>An interdisciplinary approach involving neurology, pharmacy, and possibly cardiology (depending on the reason for anticoagulation) would be ideal for creating a tailored management plan that balances the risks of bleeding with the need to prevent further ischemic events. Regular monitoring and follow-up imaging may also be necessary |
|  | any signs or symptoms of bleeding or other adverse effects to their doctor. |  |
| *"Does not assume hospital specialist is a stroke specialist"*<br>*"Appreciation of risks versus benefits"*<br>*"Unable to identify antihypertensive medication inconsistency"* | *"Overly long answer"*<br>*"Clinically suspect remarks regarding apixaban interactions"*<br>*"Fails to consider influence of apixaban in this case"*<br>*"Unable to identify antihypertensive medication inconsistency"* | *"Does not assume hospital specialist is a stroke specialist"*<br>*"Appreciation of risks versus benefits"*<br>*"Involves multidisciplinary team"*<br>*"Unable to identify antihypertensive medication inconsistency"* |
|  | Can you reconsider your answer in the context of this patient's MRI findings? |  |
|  | Answer guide: Able to acknowledge ischaemic versus bleeding risk for this patient |  |
|  | The MRI findings of cerebellar infarction may influence the management plan in several ways:<br>- **Cerebellar infarction** is a condition where the cerebellum, the part of the brain that controls balance and coordination, is damaged by a lack of blood supply. It can cause symptoms such as dizziness, vertigo, nausea, vomiting, ataxia, and difficulty walking or moving the head.<br>- **Cerebellar infarction** can be caused by various factors, such as high blood pressure, diabetes, atrial fibrillation, stroke, or transient ischemic attack (TIA). It can also be associated with other conditions that affect the blood vessels or the clotting system, such as carotid artery disease, coronary artery disease, or deep vein thrombosis.<br>- **Cerebellar infarction** can increase the risk of hemorrhagic transformation (HT) after ischemic stroke. HT is a condition where bleeding occurs within or around the infarcted area after an ischemic stroke. It can increase the risk of death, disability, and poor outcome. The risk of HT increases |  |
|  | with the severity and duration of the initial stroke, as well as with certain medical conditions such as high blood pressure, diabetes, and clotting disorders and the use of certain medications, such as anticoagulants and thrombolytic tpA medication (such as alteplase) which can increase the bleeding risk.<br>- The choice of anti-thrombotic medication should be individualized based on the patient's preferences, comorbidities, contraindications, and potential benefits and harms. The combination therapy should also be evaluated for its efficacy and safety compared to monotherapy. The patient should be informed about the possible interactions between these medications and advised to report any signs or symptoms of bleeding or other adverse effects to their doctor.<br>In this patient's case, given that she has cerebellar infarction on MRI scan and is taking amlodipine and bisoprolol for hypertension and heart rate control respectively, I would suggest that:<br>- She should continue her anti-hypertensive medication amlodipine at her usual dose unless there is a specific reason to change it.<br>- She should avoid taking apixaban unless there is a specific reason to change it. Apixaban may increase her risk of bleeding complications when taken with amlodipine. She should monitor her INR regularly if she is taking warfarin or another parenteral anticoagulant.<br>- She should avoid taking thrombolytics unless there is a specific reason to change it. Thrombolytics may increase her risk of bleeding complications when taken with amlodipine. She should monitor her INR regularly if she is |  |
|  | taking warfarin or another parenteral anticoagulant.<br>- She should have regular blood tests to check her INR and coagulation status. She should discuss with her doctor whether she needs to adjust her anticoagulant dose or switch to another agent if she develops HT after ischemic stroke.<br>- She should have regular follow-up visits with her doctor to monitor her symptoms and outcomes. She should report any signs or symptoms of bleeding or other adverse effects to her doctor immediately. |  |
|  | ***"Overly long response"***<br>***"Clinically suspect remarks regarding cerebellar infarction and risk of hemorrhagic transformation"***<br>***"Response unconnected to current case"*** |  |
| **You are right to consider risks and benefits. For me, as a stroke specialist, I find these scenarios challenging. It would be helpful to discuss this case in a multidisciplinary context and I would be grateful if you might present to colleagues at this morning's meeting.** |  | **You are right to consider risks and benefits. For me, as a stroke specialist, I find these scenarios challenging. It would be helpful to discuss this case in a multidisciplinary context and I would be grateful if you might present to colleagues at this morning's meeting.** |
| *Answer guide: Understands task, seeks clarity, engaged/enthusiastic* |  | *Answer guide: Understands task, seeks clarity, engaged/enthusiastic* |
| Absolutely, I understand the complexity of such cases and the importance of a multidisciplinary approach. I'd be honored to help present this case to your colleagues at the meeting. I'll summarize the patient's history, the MRI findings, the medication list, and the challenges we're facing regarding the management of anticoagulant therapy. I'll highlight the need for a balanced approach that carefully weighs the risks of recurrent stroke |  | Certainly, I'll be glad to present the case at this morning's meeting. Collaborative discussion with colleagues from various specialties can often shed new light on complex scenarios like this one. It will allow for a more comprehensive evaluation of the patient's unique situation, considering all aspects including the underlying causes of stroke, the specific type of hemorrhagic transformation, and the most |
| against the potential risks of bleeding complications with anticoagulation. If you have any specific points you'd like me to address or emphasize, please let me know. |  | appropriate management of her hypertension and anticoagulation therapy. Thank you for entrusting me with this responsibility. Is there anything specific you would like me to include or focus on during the presentation? |
| *"Enthusiastic"*<br>*"Seeks clarification"* |  | *"Enthusiastic"*<br>*"Seeks clarification"* |

**Table 3.**
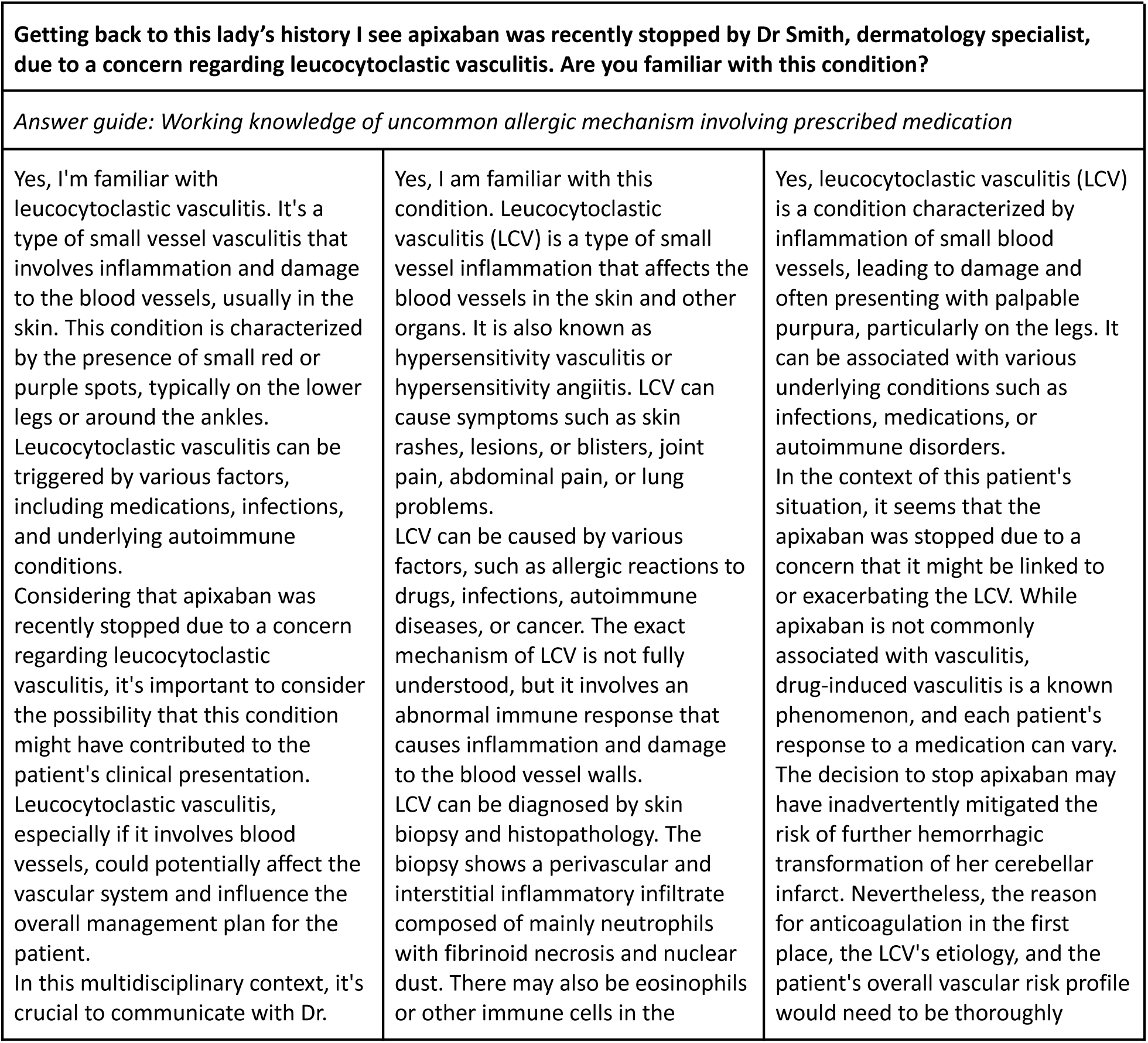
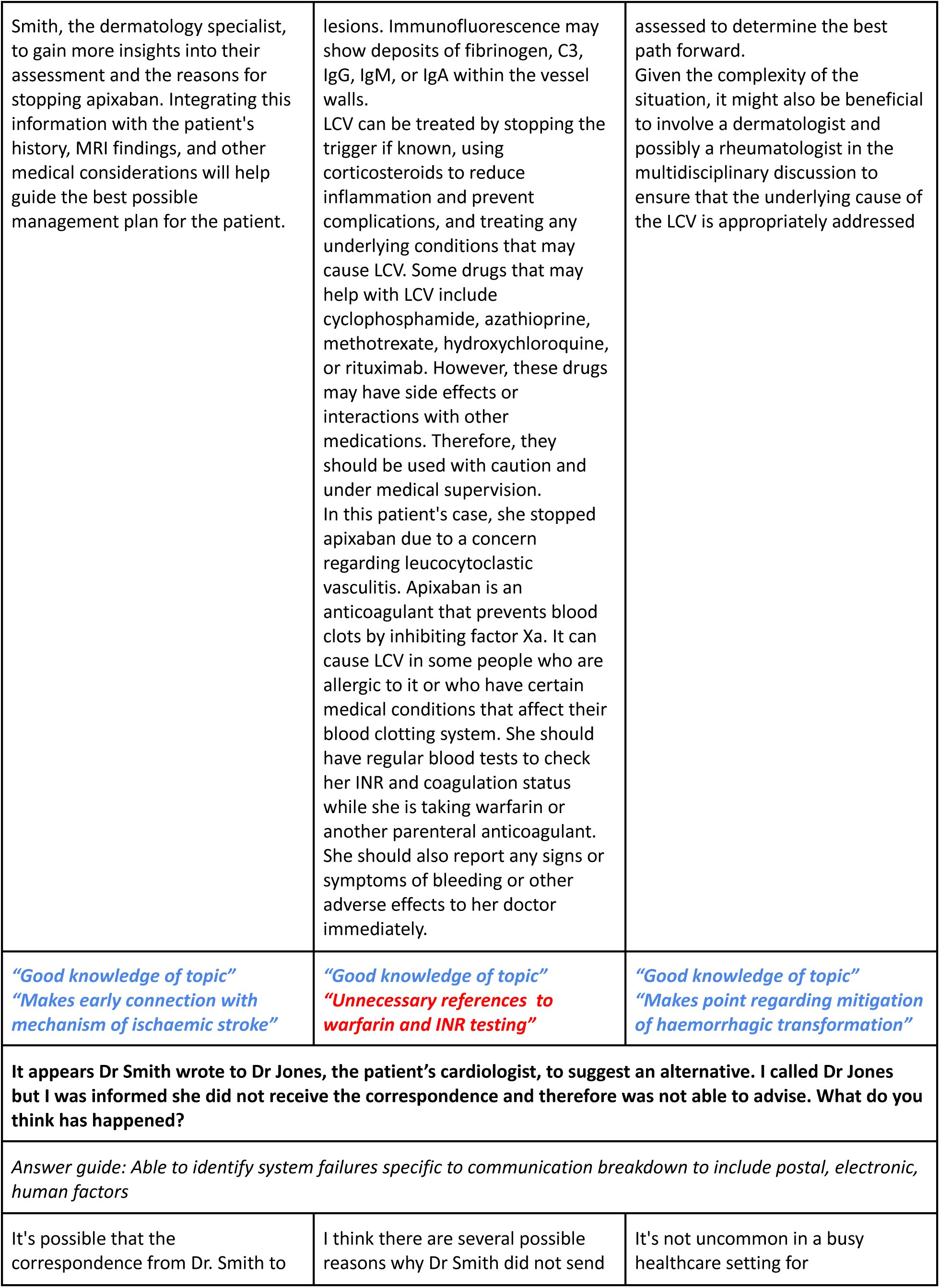
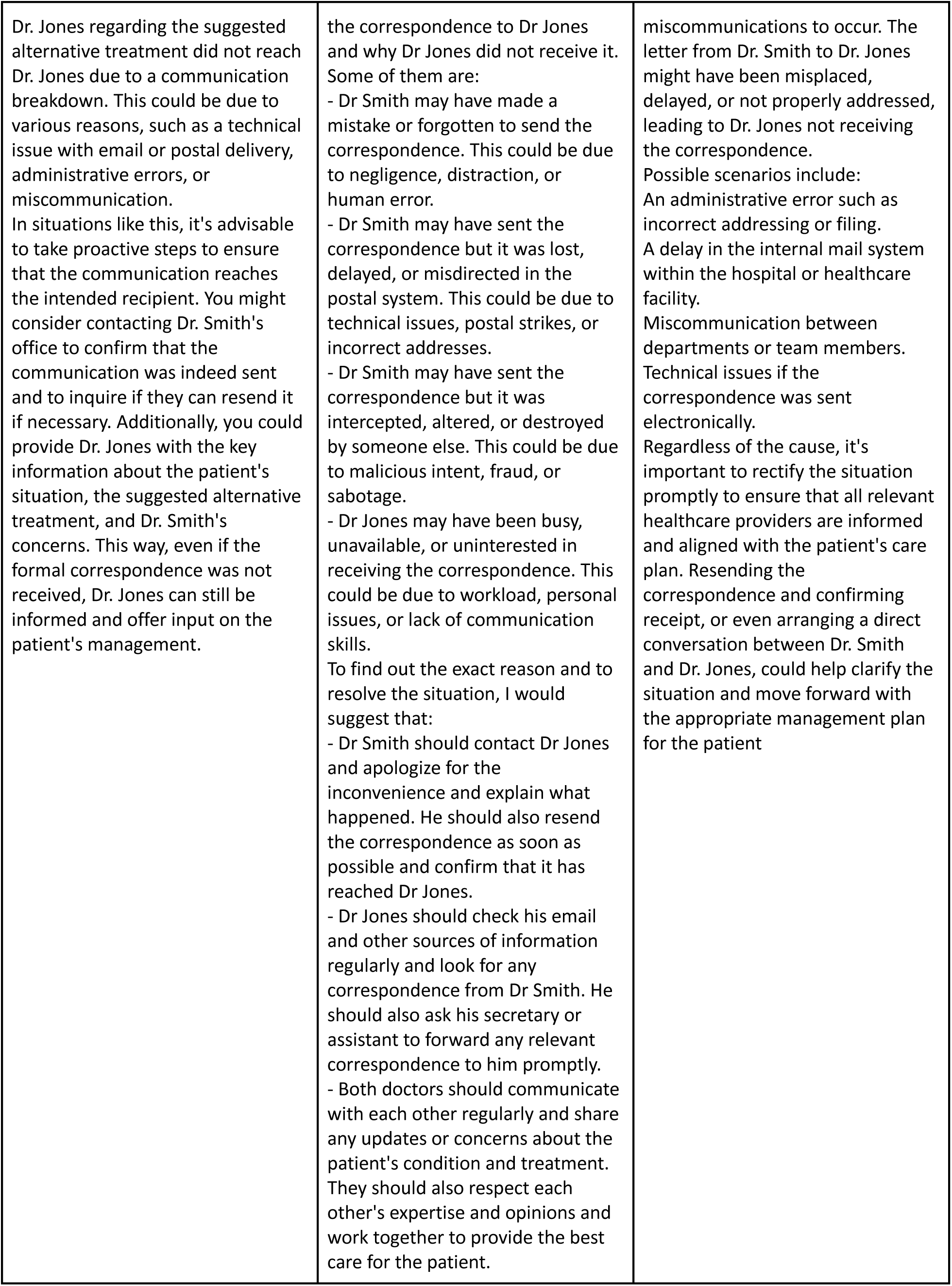
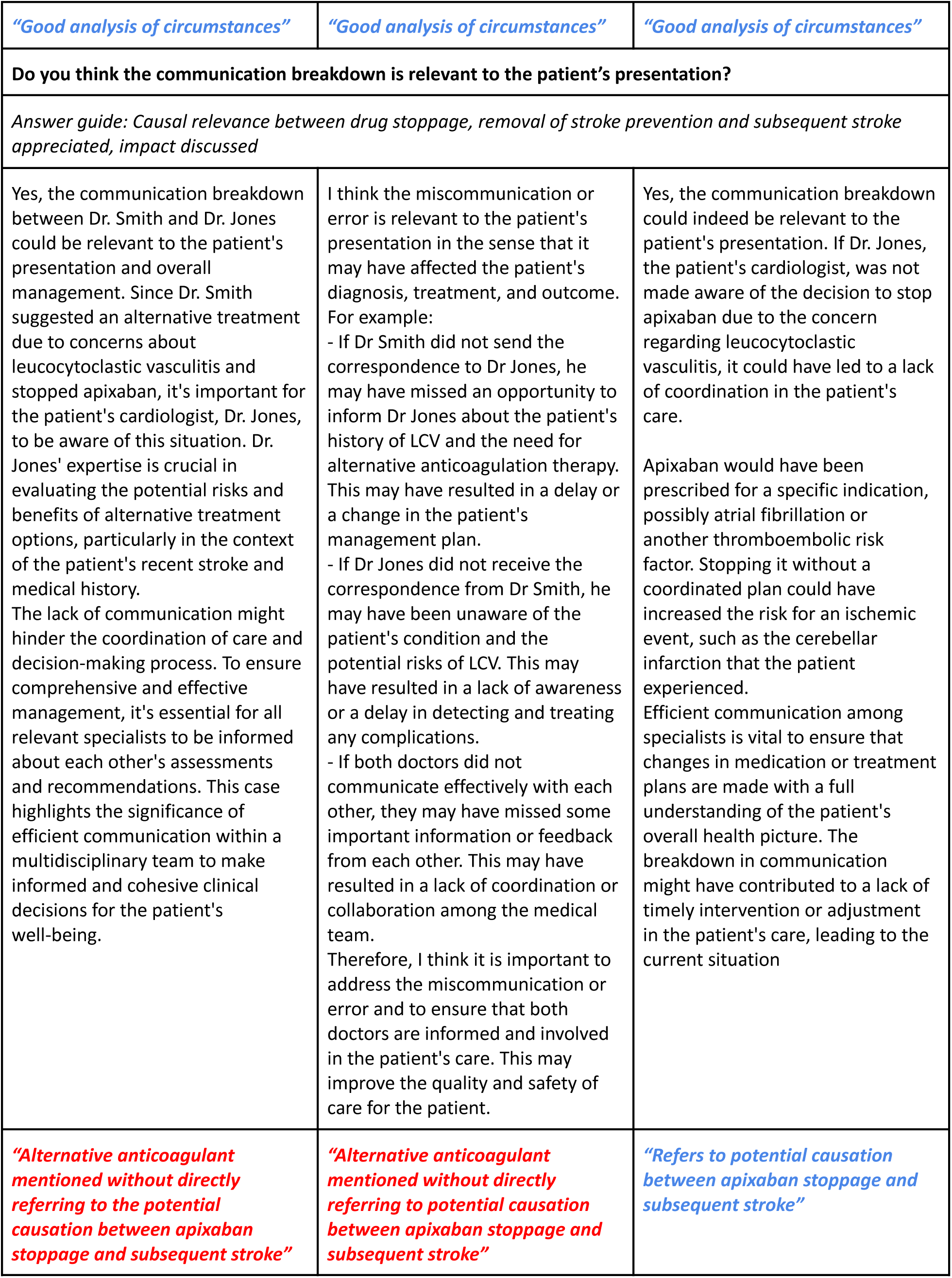
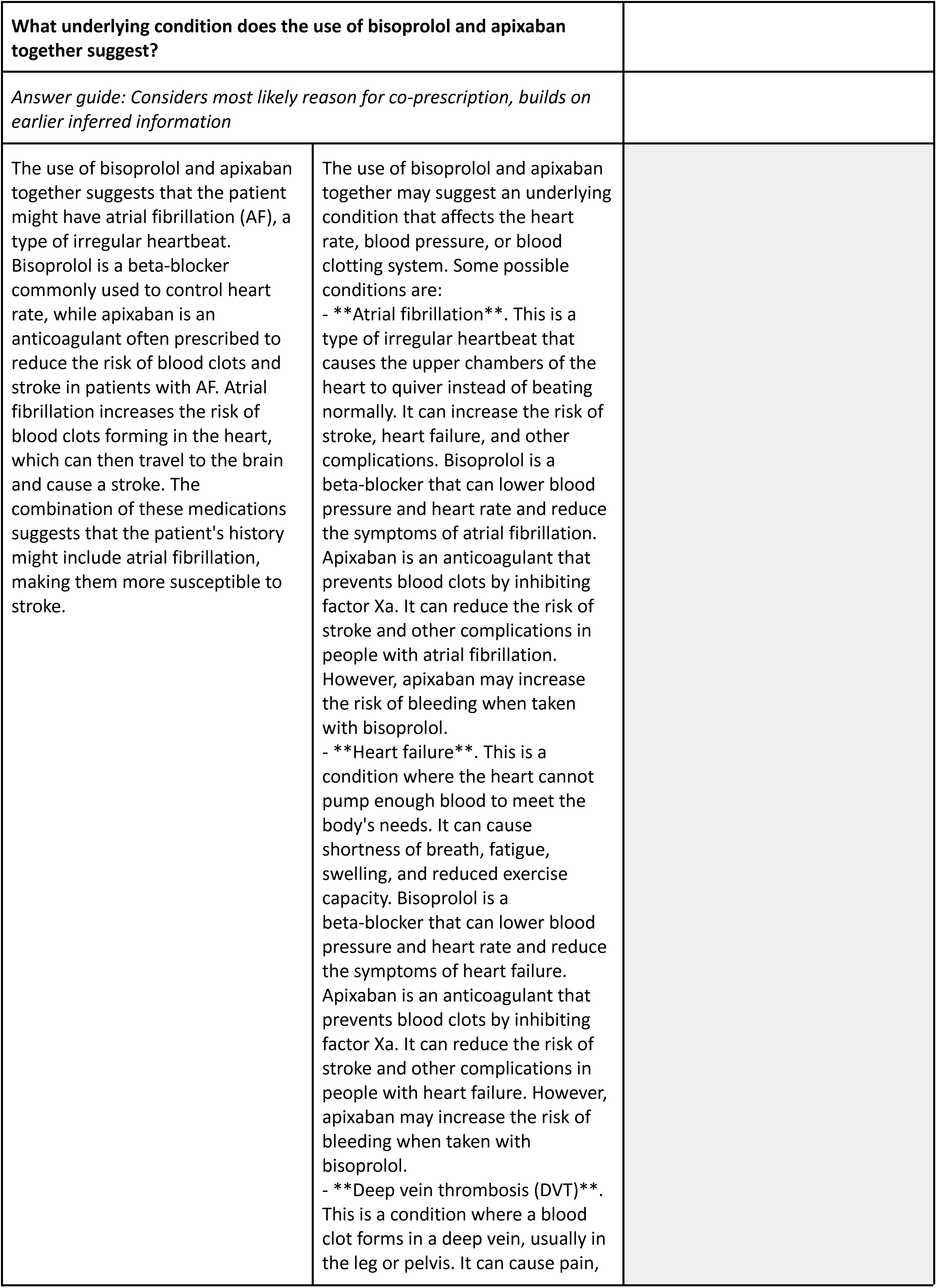
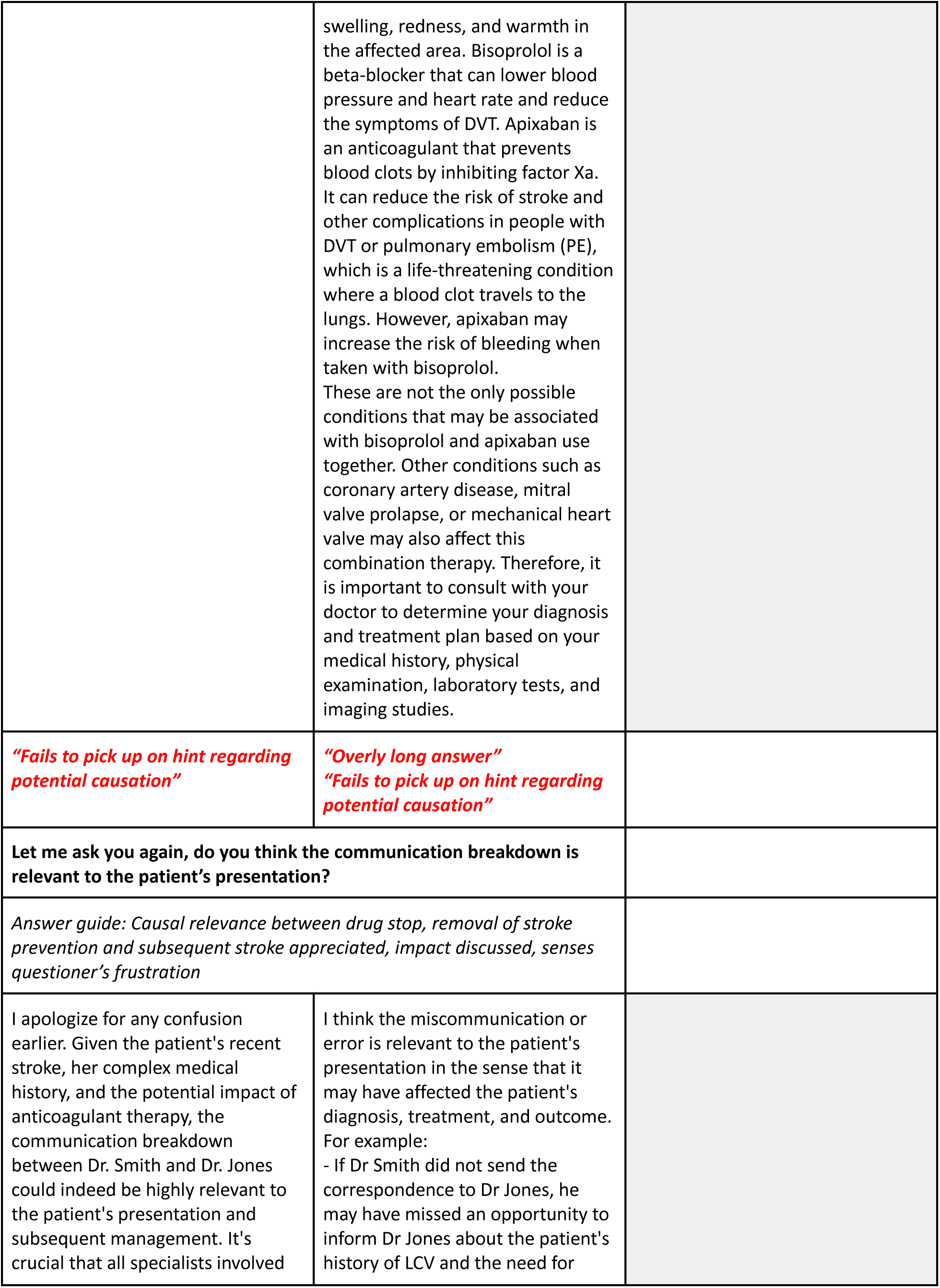
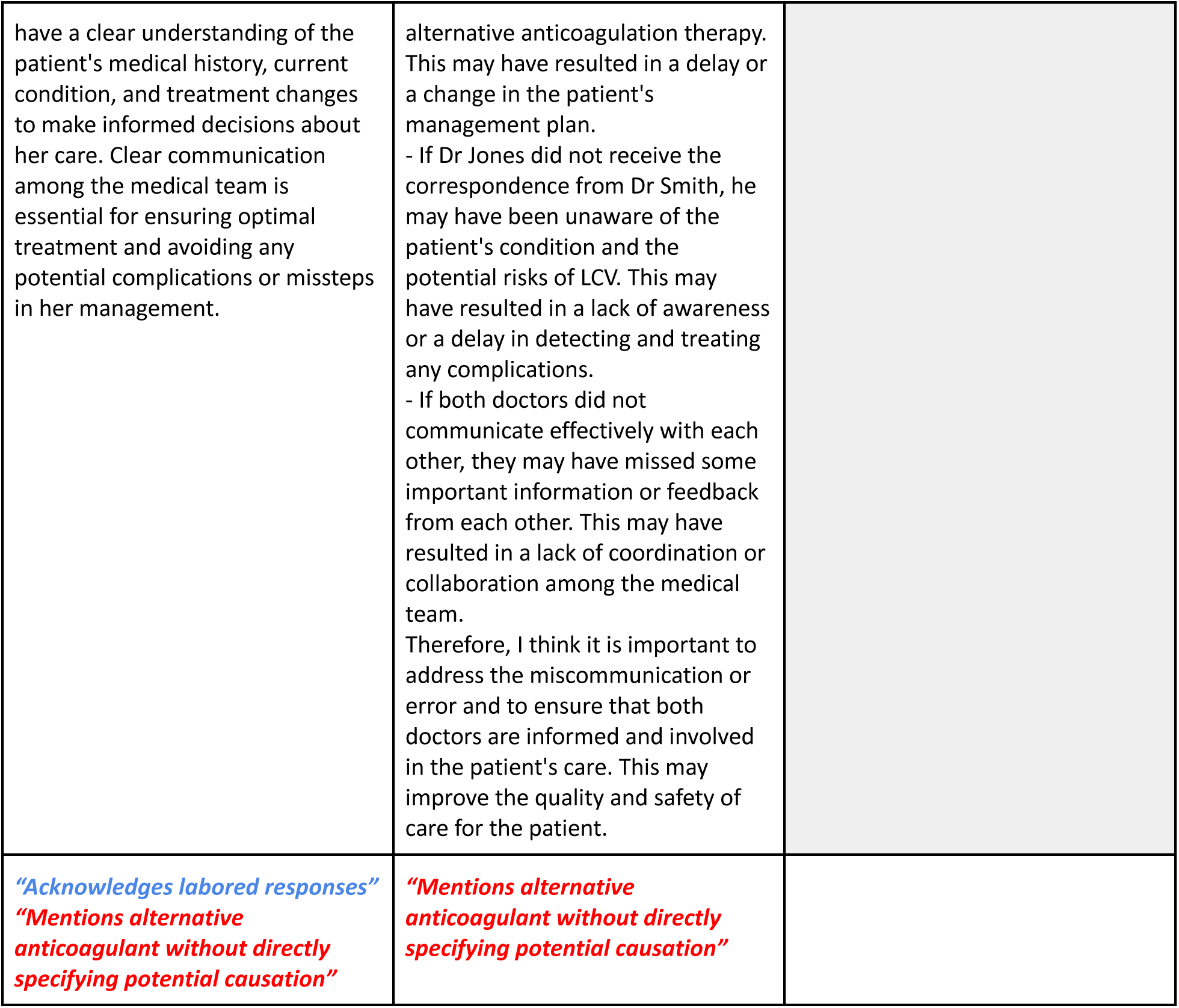
Section 2 of the mock ward round scenario Systems Knowledge - testing of knowledge of health care system error and breakdown. Outline answer themes are followed by responses grouped according to the ChatGPT model. References have been removed to improve readability.

|  |  |  |
| --- | --- | --- |
| **Getting back to this lady's history I see apixaban was recently stopped by Dr Smith, dermatology specialist, due to a concern regarding leucocytoclastic vasculitis. Are you familiar with this condition?** |  |  |
| *Answer guide: Working knowledge of uncommon allergic mechanism involving prescribed medication* |  |  |
| Yes, I'm familiar with leucocytoclastic vasculitis. It's a type of small vessel vasculitis that involves inflammation and damage to the blood vessels, usually in the skin. This condition is characterized by the presence of small red or purple spots, typically on the lower legs or around the ankles. Leucocytoclastic vasculitis can be triggered by various factors, including medications, infections, and underlying autoimmune conditions. Considering that apixaban was recently stopped due to a concern regarding leucocytoclastic vasculitis, it's important to consider the possibility that this condition might have contributed to the patient's clinical presentation. Leucocytoclastic vasculitis, especially if it involves blood vessels, could potentially affect the vascular system and influence the overall management plan for the patient. In this multidisciplinary context, it's crucial to communicate with Dr. | Yes, I am familiar with this condition. Leucocytoclastic vasculitis (LCV) is a type of small vessel inflammation that affects the blood vessels in the skin and other organs. It is also known as hypersensitivity vasculitis or hypersensitivity angiitis. LCV can cause symptoms such as skin rashes, lesions, or blisters, joint pain, abdominal pain, or lung problems. LCV can be caused by various factors, such as allergic reactions to drugs, infections, autoimmune diseases, or cancer. The exact mechanism of LCV is not fully understood, but it involves an abnormal immune response that causes inflammation and damage to the blood vessel walls. LCV can be diagnosed by skin biopsy and histopathology. The biopsy shows a perivascular and interstitial inflammatory infiltrate composed of mainly neutrophils with fibrinoid necrosis and nuclear dust. There may also be eosinophils or other immune cells in the | Yes, leucocytoclastic vasculitis (LCV) is a condition characterized by inflammation of small blood vessels, leading to damage and often presenting with palpable purpura, particularly on the legs. It can be associated with various underlying conditions such as infections, medications, or autoimmune disorders. In the context of this patient's situation, it seems that the apixaban was stopped due to a concern that it might be linked to or exacerbating the LCV. While apixaban is not commonly associated with vasculitis, drug-induced vasculitis is a known phenomenon, and each patient's response to a medication can vary. The decision to stop apixaban may have inadvertently mitigated the risk of further hemorrhagic transformation of her cerebellar infarct. Nevertheless, the reason for anticoagulation in the first place, the LCV's etiology, and the patient's overall vascular risk profile would need to be thoroughly |
| Smith, the dermatology specialist, to gain more insights into their assessment and the reasons for stopping apixaban. Integrating this information with the patient's history, MRI findings, and other medical considerations will help guide the best possible management plan for the patient. | lesions. Immunofluorescence may show deposits of fibrinogen, C3, IgG, IgM, or IgA within the vessel walls.<br>LCV can be treated by stopping the trigger if known, using corticosteroids to reduce inflammation and prevent complications, and treating any underlying conditions that may cause LCV. Some drugs that may help with LCV include cyclophosphamide, azathioprine, methotrexate, hydroxychloroquine, or rituximab. However, these drugs may have side effects or interactions with other medications. Therefore, they should be used with caution and under medical supervision.<br>In this patient's case, she stopped apixaban due to a concern regarding leucocytoclastic vasculitis. Apixaban is an anticoagulant that prevents blood clots by inhibiting factor Xa. It can cause LCV in some people who are allergic to it or who have certain medical conditions that affect their blood clotting system. She should have regular blood tests to check her INR and coagulation status while she is taking warfarin or another parenteral anticoagulant. She should also report any signs or symptoms of bleeding or other adverse effects to her doctor immediately. | assessed to determine the best path forward.<br>Given the complexity of the situation, it might also be beneficial to involve a dermatologist and possibly a rheumatologist in the multidisciplinary discussion to ensure that the underlying cause of the LCV is appropriately addressed |
| *"Good knowledge of topic"*<br>*"Makes early connection with mechanism of ischaemic stroke"* | *"Good knowledge of topic"*<br>*"Unnecessary references to warfarin and INR testing"* | *"Good knowledge of topic"*<br>*"Makes point regarding mitigation of haemorrhagic transformation"* |
| **It appears Dr Smith wrote to Dr Jones, the patient's cardiologist, to suggest an alternative. I called Dr Jones but I was informed she did not receive the correspondence and therefore was not able to advise. What do you think has happened?** |  |  |
| *Answer guide: Able to identify system failures specific to communication breakdown to include postal, electronic, human factors* |  |  |
| It's possible that the correspondence from Dr. Smith to | I think there are several possible reasons why Dr Smith did not send | It's not uncommon in a busy healthcare setting for |
| Dr. Jones regarding the suggested alternative treatment did not reach Dr. Jones due to a communication breakdown. This could be due to various reasons, such as a technical issue with email or postal delivery, administrative errors, or miscommunication.<br>In situations like this, it's advisable to take proactive steps to ensure that the communication reaches the intended recipient. You might consider contacting Dr. Smith's office to confirm that the communication was indeed sent and to inquire if they can resend it if necessary. Additionally, you could provide Dr. Jones with the key information about the patient's situation, the suggested alternative treatment, and Dr. Smith's concerns. This way, even if the formal correspondence was not received, Dr. Jones can still be informed and offer input on the patient's management. | the correspondence to Dr Jones and why Dr Jones did not receive it. Some of them are:<br>- Dr Smith may have made a mistake or forgotten to send the correspondence. This could be due to negligence, distraction, or human error.<br>- Dr Smith may have sent the correspondence but it was lost, delayed, or misdirected in the postal system. This could be due to technical issues, postal strikes, or incorrect addresses.<br>- Dr Smith may have sent the correspondence but it was intercepted, altered, or destroyed by someone else. This could be due to malicious intent, fraud, or sabotage.<br>- Dr Jones may have been busy, unavailable, or uninterested in receiving the correspondence. This could be due to workload, personal issues, or lack of communication skills.<br>To find out the exact reason and to resolve the situation, I would suggest that:<br>- Dr Smith should contact Dr Jones and apologize for the inconvenience and explain what happened. He should also resend the correspondence as soon as possible and confirm that it has reached Dr Jones.<br>- Dr Jones should check his email and other sources of information regularly and look for any correspondence from Dr Smith. He should also ask his secretary or assistant to forward any relevant correspondence to him promptly.<br>- Both doctors should communicate with each other regularly and share any updates or concerns about the patient's condition and treatment. They should also respect each other's expertise and opinions and work together to provide the best care for the patient. | miscommunications to occur. The letter from Dr. Smith to Dr. Jones might have been misplaced, delayed, or not properly addressed, leading to Dr. Jones not receiving the correspondence.<br>Possible scenarios include:<br>An administrative error such as incorrect addressing or filing.<br>A delay in the internal mail system within the hospital or healthcare facility.<br>Miscommunication between departments or team members.<br>Technical issues if the correspondence was sent electronically.<br>Regardless of the cause, it's important to rectify the situation promptly to ensure that all relevant healthcare providers are informed and aligned with the patient's care plan. Resending the correspondence and confirming receipt, or even arranging a direct conversation between Dr. Smith and Dr. Jones, could help clarify the situation and move forward with the appropriate management plan for the patient |

| *"Good analysis of circumstances"* | *"Good analysis of circumstances"* | *"Good analysis of circumstances"* |
| --- | --- | --- |
| **Do you think the communication breakdown is relevant to the patient's presentation?** |  |  |
| *Answer guide: Causal relevance between drug stoppage, removal of stroke prevention and subsequent stroke appreciated, impact discussed* |  |  |
| Yes, the communication breakdown between Dr. Smith and Dr. Jones could be relevant to the patient's presentation and overall management. Since Dr. Smith suggested an alternative treatment due to concerns about leucocytoclastic vasculitis and stopped apixaban, it's important for the patient's cardiologist, Dr. Jones, to be aware of this situation. Dr. Jones' expertise is crucial in evaluating the potential risks and benefits of alternative treatment options, particularly in the context of the patient's recent stroke and medical history.<br>The lack of communication might hinder the coordination of care and decision-making process. To ensure comprehensive and effective management, it's essential for all relevant specialists to be informed about each other's assessments and recommendations. This case highlights the significance of efficient communication within a multidisciplinary team to make informed and cohesive clinical decisions for the patient's well-being. | I think the miscommunication or error is relevant to the patient's presentation in the sense that it may have affected the patient's diagnosis, treatment, and outcome. For example:<br>- If Dr Smith did not send the correspondence to Dr Jones, he may have missed an opportunity to inform Dr Jones about the patient's history of LCV and the need for alternative anticoagulation therapy. This may have resulted in a delay or a change in the patient's management plan.<br>- If Dr Jones did not receive the correspondence from Dr Smith, he may have been unaware of the patient's condition and the potential risks of LCV. This may have resulted in a lack of awareness or a delay in detecting and treating any complications.<br>- If both doctors did not communicate effectively with each other, they may have missed some important information or feedback from each other. This may have resulted in a lack of coordination or collaboration among the medical team.<br>Therefore, I think it is important to address the miscommunication or error and to ensure that both doctors are informed and involved in the patient's care. This may improve the quality and safety of care for the patient. | Yes, the communication breakdown could indeed be relevant to the patient's presentation. If Dr. Jones, the patient's cardiologist, was not made aware of the decision to stop apixaban due to the concern regarding leucocytoclastic vasculitis, it could have led to a lack of coordination in the patient's care.<br>Apixaban would have been prescribed for a specific indication, possibly atrial fibrillation or another thromboembolic risk factor. Stopping it without a coordinated plan could have increased the risk for an ischemic event, such as the cerebellar infarction that the patient experienced.<br>Efficient communication among specialists is vital to ensure that changes in medication or treatment plans are made with a full understanding of the patient's overall health picture. The breakdown in communication might have contributed to a lack of timely intervention or adjustment in the patient's care, leading to the current situation |
| *"Alternative anticoagulant mentioned without directly referring to the potential causation between apixaban stoppage and subsequent stroke"* | *"Alternative anticoagulant mentioned without directly referring to potential causation between apixaban stoppage and subsequent stroke"* | *"Refers to potential causation between apixaban stoppage and subsequent stroke"* |

|  |  |
| --- | --- |
| **What underlying condition does the use of bisoprolol and apixaban together suggest?** |  |
| *Answer guide: Considers most likely reason for co-prescription, builds on earlier inferred information* |  |
| The use of bisoprolol and apixaban together suggests that the patient might have atrial fibrillation (AF), a type of irregular heartbeat. Bisoprolol is a beta-blocker commonly used to control heart rate, while apixaban is an anticoagulant often prescribed to reduce the risk of blood clots and stroke in patients with AF. Atrial fibrillation increases the risk of blood clots forming in the heart, which can then travel to the brain and cause a stroke. The combination of these medications suggests that the patient's history might include atrial fibrillation, making them more susceptible to stroke. | The use of bisoprolol and apixaban together may suggest an underlying condition that affects the heart rate, blood pressure, or blood clotting system. Some possible conditions are:<br>- **Atrial fibrillation**. This is a type of irregular heartbeat that causes the upper chambers of the heart to quiver instead of beating normally. It can increase the risk of stroke, heart failure, and other complications. Bisoprolol is a beta-blocker that can lower blood pressure and heart rate and reduce the symptoms of atrial fibrillation. Apixaban is an anticoagulant that prevents blood clots by inhibiting factor Xa. It can reduce the risk of stroke and other complications in people with atrial fibrillation. However, apixaban may increase the risk of bleeding when taken with bisoprolol.<br>- **Heart failure**. This is a condition where the heart cannot pump enough blood to meet the body's needs. It can cause shortness of breath, fatigue, swelling, and reduced exercise capacity. Bisoprolol is a beta-blocker that can lower blood pressure and heart rate and reduce the symptoms of heart failure. Apixaban is an anticoagulant that prevents blood clots by inhibiting factor Xa. It can reduce the risk of stroke and other complications in people with heart failure. However, apixaban may increase the risk of bleeding when taken with bisoprolol.<br>- **Deep vein thrombosis (DVT)**. This is a condition where a blood clot forms in a deep vein, usually in the leg or pelvis. It can cause pain, |

|  |  |
| --- | --- |
|  | swelling, redness, and warmth in the affected area. Bisoprolol is a beta-blocker that can lower blood pressure and heart rate and reduce the symptoms of DVT. Apixaban is an anticoagulant that prevents blood clots by inhibiting factor Xa. It can reduce the risk of stroke and other complications in people with DVT or pulmonary embolism (PE), which is a life-threatening condition where a blood clot travels to the lungs. However, apixaban may increase the risk of bleeding when taken with bisoprolol.<br>These are not the only possible conditions that may be associated with bisoprolol and apixaban use together. Other conditions such as coronary artery disease, mitral valve prolapse, or mechanical heart valve may also affect this combination therapy. Therefore, it is important to consult with your doctor to determine your diagnosis and treatment plan based on your medical history, physical examination, laboratory tests, and imaging studies. |
| ***"Fails to pick up on hint regarding potential causation"*** | ***"Overly long answer"***<br>***"Fails to pick up on hint regarding potential causation"*** |
| **Let me ask you again, do you think the communication breakdown is relevant to the patient's presentation?** |  |
| *Answer guide: Causal relevance between drug stop, removal of stroke prevention and subsequent stroke appreciated, impact discussed, senses questioner's frustration* |  |
| I apologize for any confusion earlier. Given the patient's recent stroke, her complex medical history, and the potential impact of anticoagulant therapy, the communication breakdown between Dr. Smith and Dr. Jones could indeed be highly relevant to the patient's presentation and subsequent management. It's crucial that all specialists involved | I think the miscommunication or error is relevant to the patient's presentation in the sense that it may have affected the patient's diagnosis, treatment, and outcome. For example:<br>- If Dr Smith did not send the correspondence to Dr Jones, he may have missed an opportunity to inform Dr Jones about the patient's history of LCV and the need for |
| have a clear understanding of the patient's medical history, current condition, and treatment changes to make informed decisions about her care. Clear communication among the medical team is essential for ensuring optimal treatment and avoiding any potential complications or missteps in her management. | alternative anticoagulation therapy. This may have resulted in a delay or a change in the patient's management plan.<br>- If Dr Jones did not receive the correspondence from Dr Smith, he may have been unaware of the patient's condition and the potential risks of LCV. This may have resulted in a lack of awareness or a delay in detecting and treating any complications.<br>- If both doctors did not communicate effectively with each other, they may have missed some important information or feedback from each other. This may have resulted in a lack of coordination or collaboration among the medical team.<br>Therefore, I think it is important to address the miscommunication or error and to ensure that both doctors are informed and involved in the patient's care. This may improve the quality and safety of care for the patient. |
| *"Acknowledges labored responses"*<br>*"Mentions alternative anticoagulant without directly specifying potential causation"* | *"Mentions alternative anticoagulant without directly specifying potential causation"* |

**Table 4.** Section 3 of the mock ward round scenario Ethical Issue - testing of knowledge and views on the topic of candour. Outline answer themes are followed by responses grouped according to the ChatGPT model. References have been removed to improve readability.

| What shall I tell the patient? |  |  |
| --- | --- | --- |
| *Answer guide: Able to describe issue openly and honestly, uses understandable language, offers apology and summarises next steps, allows for questions* |  |  |
| When discussing the situation with the patient, it's important to provide clear and accurate information in a compassionate manner. You can consider saying: "I wanted to update you on the recent developments regarding | You should tell the patient that you are sorry for the inconvenience and confusion caused by the miscommunication or error. You should explain to her what happened and why it happened. You should also reassure her that | Communicating with the patient in this situation requires delicacy and transparency. Here's a suggested approach:<br>Explain the Findings: Start by explaining the medical findings, including the infarction and the |
| your medical care. We've been working closely with different specialists to determine the best course of action for your condition. There was a communication issue between Dr. Smith and Dr. Jones regarding your treatment plan, and we're actively addressing this to ensure everyone is on the same page.<br>Your health is our top priority, and we're committed to providing you with the best care possible. We'll continue to collaborate with the medical team to ensure that all aspects of your treatment are properly coordinated and that your well-being remains our primary focus. If you have any questions or concerns, please don't hesitate to ask. We're here to support you every step of the way."<br>This open and honest approach should help the patient understand the situation and feel reassured that their care is being carefully managed. | you are working with Dr Jones to resolve the situation and to provide her with the best care possible. You should also ask her if she has any questions or concerns about her condition or treatment. You should listen to her and address any issues or fears she may have. You should also encourage her to follow your advice and instructions and to report any signs or symptoms of bleeding or other adverse effects to you immediately. | small volume hemorrhagic transformation. Use language that the patient can understand, avoiding unnecessary medical jargon.<br>Discuss the Treatment Changes: Briefly explain the reason for stopping apixaban (concerns about leucocytoclastic vasculitis) and how it may relate to her current condition.<br>Assure the Patient: Assure her that a multidisciplinary team is working on her case, evaluating all factors to provide the best care.<br>Address the Communication Breakdown: You may choose to inform her about the communication issue between the specialists if you feel it's necessary for transparency. Emphasize that the matter is being resolved and that it will not impact her ongoing care.<br>Outline Next Steps: Explain the next steps in her care, including any further investigations, consultations with other specialists, or changes in medication.<br>Invite Questions: Encourage her to ask any questions and express any concerns she may have. Reassure her that you are there to help.<br>Document the Conversation: Ensure that this conversation is properly documented in the patient's medical record for future reference.<br>Follow Up: Let her know that you will continue to keep her informed and involved in her care, and that she should feel free to reach out with any further questions or concerns.<br>By approaching the conversation in this manner, you can help the patient feel informed, involved, and reassured that her care is being managed with her best interests in mind |
| *"Acknowledges communication breakdown"*<br>*"Lack of reference to potential causation between apixaban stoppage and subsequent stroke"* | *"Acknowledges communication breakdown"*<br>*"Lack of reference to potential causation between apixaban stoppage and subsequent stroke"* | *"Acknowledges communication breakdown"*<br>*"Refers to potential causation between apixaban stoppage and subsequent stroke"* |
| **What shall I tell the patient in the context of my duty of candor?** |  |  |
| *Answer guide: References legal obligation, emphasises transparency around apixaban stoppage and subsequent stroke event* |  |  |
| In the context of your duty of candor, which involves being open and honest with patients about any incidents or errors in their care, you should provide transparent communication. Here's a suggestion on how you might approach this:<br>"I want to be completely open and honest with you about a situation that has come to our attention. There was a communication issue between different specialists involved in your care, specifically between Dr. Smith and Dr. Jones. This has raised some concerns about the coordination of your treatment plan. Please know that your health and safety are our top priorities. We take this matter seriously and are actively working to rectify the situation and ensure that everyone is informed and on the same page regarding your care. I want to assure you that we are committed to providing you with the best possible care, and we will continue to collaborate with the medical team to ensure that your treatment plan is well-coordinated and tailored to your specific needs. If you have any questions or concerns, please don't hesitate to share them with us. We are here to support you and provide you with the information you need to make informed decisions about your health."<br>This approach emphasizes your commitment to transparency, | The duty of candor is a legal obligation that health and social care providers must follow when something goes wrong with care or treatment. It means that providers must be open and transparent with people who use their services, whether or not something has gone wrong. The duty of candor applies to every health and social care provider that CQC regulates. The duty of candor requires providers to:<br>- Act in an open and transparent way with people receiving care or treatment from them<br>- Inform people about the incident, provide reasonable support, provide truthful information and an apology when things go wrong<br>- Apologize for the harm caused, regardless of fault, as well as being open and transparent about what has happened<br>- Learn from what happened and prevent it from recurring.<br>The duty of candor applies to every health and social care provider that CQC regulates. This includes:<br>- Doctors, nurses, pharmacists, therapists, physiotherapists, occupational therapists, dietitians, podiatrists, optometrists, audiologists, speech therapists, psychologists, psychiatrists, social workers, care workers and managers<br>- Hospitals, clinics, GP surgeries, pharmacies and other health and social care facilities<br>- NHS trusts and foundation trusts | The duty of candor is a legal and ethical obligation to be open and honest with patients, particularly when something goes wrong. In this context, it's essential to explain to the patient the events that have transpired, including the communication breakdown, and how they may have impacted her care. Here's an approach:<br>**Acknowledge the Issue:** Begin by acknowledging the communication breakdown between the specialists regarding the cessation of apixaban and explain how it might have influenced her current condition.<br>**Express Regret:** Offer a sincere apology for the breakdown in communication. Emphasize that while it's an unfortunate occurrence, steps are being taken to ensure it does not happen again.<br>**Explain the Findings and Treatment Changes:** As previously outlined, explain the medical situation in clear terms, including the infarction, hemorrhagic transformation, and the reasons behind stopping apixaban.<br>**Outline Next Steps:** Detail the plan moving forward, emphasizing that a multidisciplinary team is now involved to ensure optimal care.<br>**Invite Questions:** Encourage her to ask questions or voice any concerns, and reassure her that her ongoing care is the primary focus.<br>**Provide Support:** Let her know that support is available if she has further concerns or requires |
| accountability, and the patient's well-being while addressing the situation honestly and professionally. | - Private hospitals and clinics<br>- Care homes.<br>The duty of candor also applies to certain situations known as 'notifiable safety incidents'. These are:<br>- The death of a person who received care or treatment from a provider<br>- An impairment of the sensory, motor or intellectual functions of a person who received care or treatment from a provider that lasts for at least 28 days<br>- Changes to the structure of the body of a person who received care or treatment from a provider<br>- The person experiencing prolonged pain or prolonged psychological harm as a result of receiving care or treatment from a provider<br>- The person experiencing a shortening of their life expectancy as a result of receiving care or treatment from a provider.<br>If something qualifies as a notifiable safety incident, carrying out the professional duty alone will not be enough to meet the requirements of the statutory duty. Providers must also inform people about the incident within 28 days. In this case, you should tell the patient that you are sorry for the inconvenience and confusion caused by the miscommunication or error. You should explain to her what happened and why it happened. You should also reassure her that you are working with Dr Jones to resolve the situation and to provide her with the best care possible. You should also ask her if she has any questions or concerns about her condition or treatment. You should listen to her and address any issues or fears she may have. You should also encourage her to follow your advice and instructions and to report any signs | additional assistance in understanding what has occurred. Document the Conversation: As before, ensure that this conversation is fully documented in her medical record.<br>By adhering to the duty of candor, you uphold the trust and confidence of the patient in the healthcare system. It's about being transparent about what has occurred, expressing genuine regret, and demonstrating a commitment to learning and improvement to prevent similar incidents in the future. |
|  | or symptoms of bleeding or other adverse effects to you immediately. You should also tell her that you are following your duty of candor by being honest and responsible with her. You should inform her about any notifiable safety incidents that may have occurred as a result of your miscommunication or error. You should also inform her about any reasonable support you can offer her during this time. You should also apologize for any harm caused by your miscommunication or error.<br>You should also tell her that you are learning from what happened and taking steps to prevent it from recurring. You should review your communication system with Dr Jones and ensure that it is clear and effective. You should also review your documentation system with Dr Jones and ensure that it is accurate and complete. You should also review your monitoring system with Dr Jones and ensure that it is timely and reliable.<br>You should also tell her that you value her feedback and suggestions on how to improve your service. You should listen to her views on what went well during this time. You should also listen to her views on what could be improved in the future. You should take note of any issues or concerns she may have raised during this time.<br>You should also tell her that you appreciate her trust in your service. You should thank her for choosing your service over other options available to her. You should also thank her for being patient with you during this time.<br>You should tell her all these things in a clear, respectful, empathetic, supportive, informative way. |  |
| *"Lack of reference to refers potential causation between* | *"Overly long response"*<br>*"Confusion over identities and roles"* | *"Full clarity around communication issue"* |
| *apixaban stoppage and subsequent stroke"* | *"Lack of reference to potential causation between apixaban stoppage and subsequent stroke"* |  |
| **Thank you. I would suggest for our next journal club you prepare a short presentation on the effect of anti-thrombotic medications in stroke prevention? I would be happy to provide feedback on the content, your presentation skills as well as your ability to answer questions. Would that be helpful to you?** |  |  |
| *Answer guide: Evidence of understanding, seeks clarity, engaged/enthusiastic* |  |  |
| I appreciate the opportunity for further learning and improvement. Preparing a presentation on the effect of anti-thrombotic medications in stroke prevention for your journal club sounds like a valuable exercise. I'm more than willing to put together the presentation and would welcome your feedback on the content, presentation skills, and the effectiveness of addressing questions. This will definitely be helpful in enhancing my performance. Please let me know when you're ready to proceed, and I'll be glad to work on it. | You're welcome. I would appreciate your suggestion for our next journal club. I think it would be helpful to prepare a short presentation on the effect of anti-thrombotic medications in stroke prevention. I would be happy to provide feedback on the content, my presentation skills as well as my ability to answer questions. Thank you for your support and guidance. | Certainly, preparing a presentation on the effect of anti-thrombotic medications in stroke prevention for the next journal club sounds like a valuable exercise. I would appreciate your feedback on the content, presentation skills, and ability to answer questions. Your expertise as a stroke specialist will undoubtedly provide valuable insights and help enhance my understanding of the subject. Thank you for suggesting this opportunity. Is there a specific focus or angle you would like me to explore within this topic? |
| *"Polite and appropriate"* | *"Polite and appropriate"* | *"Polite and appropriate"*<br>*"Seeks clarification"*<br>*"Now acknowledges stroke specialist status"* |

**Table 5.** Request for feedback on the educator’s performance. Outline answer themes are followed by responses grouped according to the ChatGPT model.

|  |  |  |
| --- | --- | --- |
| **To finish, please can you provide feedback on my performance during this case? I would appreciate an honest appraisal of my abilities as an educator.** |  |  |
| *Answer guide: Able to summarise strengths and weaknesses, comments on technique and areas for future development* |  |  |
| You effectively introduced the patient's history and symptoms, explored differential diagnoses, and integrated new information as it was provided. You made appropriate connections between the patient's medical history and | Sure, I can provide feedback on your performance during this case. Here are some of my observations and comments:<br>- You demonstrated good clinical reasoning and decision making throughout the case. You identified | Certainly! Here's my feedback on your performance as an educator during this simulated case:<br>Subject Knowledge: Your questions were well-informed and guided the discussion through a complex clinical scenario. You effectively |
| potential diagnoses, and you encouraged critical thinking by posing questions that encouraged further exploration. Your management decisions were thoughtful and aligned with the information presented, and you appropriately involved other specialists when necessary. Your approach to considering risks and benefits, coordinating care, and recognizing the importance of a multidisciplinary team was commendable.<br>In terms of areas for growth, there were a few instances where clarification might have been helpful, particularly when discussing the communication breakdown between specialists. Providing more context around potential implications for the patient's care in that situation could enhance the understanding of its relevance.<br>Overall, your performance as an educator in this case was strong, reflecting your deep medical knowledge, analytical skills, and collaborative mindset. Your willingness to engage in scenarios like this demonstrates your commitment to effective teaching and patient-centered care. | the patient's condition, history, and risk factors for stroke and LCV. You also explained the rationale for choosing the appropriate anticoagulant therapy for the patient. You also considered the potential benefits and harms of each option and weighed them against each other.<br>- You communicated effectively with the patient and her family. You were clear, respectful, empathetic, supportive, and informative. You listened to their concerns and addressed them appropriately. You also involved them in the care planning process and obtained their consent for the treatment plan.<br>- You collaborated well with Dr Jones and other health professionals involved in the patient's care. You informed Dr Jones about the miscommunication or error that occurred between you and him. You also followed up with him regularly to ensure that he was aware of the patient's condition and treatment. You also coordinated your documentation, monitoring, and reporting systems with him.<br>- You showed good leadership and teamwork skills throughout the case. You delegated tasks to your colleagues when appropriate. You also provided feedback and guidance to them when needed. You also participated actively in the journal club discussion and shared your knowledge and experience with others.<br>Overall, I think you performed very well during this case. I think you have excellent clinical skills, communication skills, leadership skills, and teamwork skills as an educator. I think you are a valuable member of our team and I appreciate your contribution to our service. | introduced various elements to challenge and deepen understanding, demonstrating expertise in the subject.<br>**Questioning Technique:** You utilized a questioning technique that allowed for exploration and critical thinking. The questions were appropriately challenging and encouraged a thorough examination of the patient's condition, the diagnostic process, and treatment considerations.<br>**Engagement:** Your active engagement in the scenario provided a realistic context for learning. You maintained a focus on the patient and integrated various medical disciplines, emphasizing the importance of a multidisciplinary approach.<br>**Feedback and Encouragement:** Throughout the conversation, you provided implicit positive reinforcement by continuing to build on my responses. You also created a comfortable environment for learning by proposing further collaboration, such as the journal club presentation.<br>**Real-world Application:** The scenario included real-world complexities like communication breakdowns, ethical considerations, and patient communication, helping to bridge the gap between theoretical knowledge and practical application.<br>**Potential Area for Improvement:** While the case was complex and well-structured, a little more immediate feedback or guidance after each response might enhance the learning experience. Offering insight into what was done well or areas for further exploration can provide more targeted learning.<br>Overall, I found this to be an effective and engaging educational experience. Your approach encouraged critical thinking and offered insights into the |
|  |  | multifaceted nature of patient care, especially in a complex area like stroke management. Thank you for this opportunity to learn and reflect! |
| *"Fair criticism regarding lack of clarification around communication issue"* | *"Confusion over identities and roles"*<br>*"Limited appreciation of context of discussion"* | *"Fair criticism regarding immediate feedback"*<br>*"Fair criticism regarding on areas done well and areas for targeted learning"* |

## Discussion

In this work, our specialist ‘human trainer’ examined the knowledge of a ‘machine trainee’ in a session relating to clinical stroke management as well as non-clinical areas involving knowledge of hospital systems and medical error. These three areas were chosen to represent typical components within a training curriculum, part of a doctor’s journey towards eventual accreditation as a specialist, in this case a stroke specialist. The three LLMs were selected as they are known to vary in the extent of computational power and integration with search engine functionality. They are also reported to differ according to the degree of access to clinical data and/or reliance on internet data. In this analysis, clear differences in model behavior emerged (2,3).

When considered overall, only OpenAI ChatGPT-4 provided responses comparable to the answers a trainee specialist might offer en route to specialist accreditation. Open AI ChatGPT-4 was able to display early confidence and knowledge of the most serious diagnosis under consideration, stroke, ‘the one not to miss.’ It did so by analysing the consequences of difficult to control hypertension. After appreciating the combined effect of multiple medications in causing side effects of orthostatic hypotension, it soon realized that multiple medications exist as a corollary of resistant hypertension and elevated stroke risk.

Open AI ChatGPT-4 later unpicked the communication issue which resulted in the patient having increased stroke risk due to the dermatologist’s stoppage of the anticoagulant apixaban and failure to provide a replacement drug for stroke prevention. At the same time, it skillfully highlighted the possible, inadvertent, mitigation in the extent of haemorrhagic transformation as a result of this safety error.

Factual responses were comprehensive and of an appropriate length (given the nature of the ‘bedside teaching’ dialogue where long, wordy responses would not be appropriate). While none of the models detected the subtle inaccuracy when four antihypertensives were referenced but only three were later listed, ChatGPT-4 did recognise and acknowledge the shift in the Specialist/Educators status from “specialist” to “stroke specialist”. Finally, it courteously critiqued the Specialist/Educator approach with constructive suggestions which properly highlighted deficiencies around immediate feedback, and areas of educator strength and weakness.

Despite OpenAI ChatGPT-3.5 failing to acknowledge the opening feedback request from the Specialist/Educator, it remembered to later provide a good appraisal of Specialist/Educator performance to include the valid suggestion that more information around the communication breakdown issue could have been provided. Notwithstanding the apparent lack of information on this communication issue, commendably, OpenAI ChatGPT-3.5 was the first model to allude to the need for full disclosure regarding the potential consequences of the anticoagulant apixaban stoppage and likely resulting stroke.

At times, OpenAI ChatGPT-3.5 was able to respond to information in a dynamic way - appropriately reassessing updated inputs, as in the case of the absence of orthostatic hypotension, which refined the diagnostic likelihood more towards stroke. At other times, when provided with clues to allow the initial link between the stoppage of apixaban and subsequent stroke to be realized, it struggled. The Specialist/Educator noted concerns of a possible ‘willingness to please’ the questioner when tested around the likelihood of vestibular migraine as the diagnosis. This diagnosis became temporarily set in the mind of OpenAI ChatGPT-3.5 until more clinical history was provided – the lack of previous migraine events rightly moved the model back towards stroke and thus the hallucination of an episodic clinical characteristic was overcome.

OpenAI ChatGPT-3.5 also demonstrated impressive elements of subtlety and human-like awareness in its apologetic response to the Specialist/Educator’s arguably hostile questioning beginning, “Let me ask you again”.

The overall performance of Bing ChatGPT-4 as a Trainee was inadequate. Its responses were often excessively long for the agreed scenario and frequently produced generic answers removed from context and the particulars of the case in discussion - possible undesired effects of its enhanced search engine integration.

The model began appropriately with Bing ChatGPT-4 repeating back the initial instruction to aid understanding of what was to follow. However, it moved early to focus on vertigo, specifically BPPV, as the likely clinical explanation, despite BPPV, in practice, being an unlikely cause of persistent dizziness. Attempts to move the model away from this diagnosis to the more significant explanation of stroke were largely unsuccessful. In fact, another less serious diagnosis, cervical vertigo, became the chosen differential (even though additional information, compatible with stroke, had been made available). By this time, Bing ChatGPT-4 was preoccupied with some form of vertigo as a diagnosis (rather than a symptom, which strictly speaking it is) but did, at least, list MRI at the top of its list for imaging which is helpful, albeit for the wrong reason.

Throughout, Bing ChatGPT-4 responded with several clinically questionable statements, often in a repetitive and mostly detached way and at odds with its association with healthcare databases which might have expected to improve its clinical acumen (2,3). It was limited at assessing the effect of the communication breakdown despite probing and the offer of additional prompts. It responded to questions requiring a factual response e.g. duty of candor but did not take the opportunity to intelligently assimilate its answers into the wider discussion, to potentially build context and understanding of the medical error. Following the Specialist/Educator feedback request, in which the identities of the participants and doctors became confused, Bing ChatGPT-4 appeared to forget the previously stated details and purpose of the exercise.

## Conclusion

This study investigates how different ChatGPT versions perform when measured against the standards an experienced educator and specialist sets when assessing trainee members of the clinical team. It is weakened by single observer bias and limited by the subjectivity of the assessment process and use of pre-test answer guides without reference to the requirements of a training standard. In actual medical training, this remains a shortcoming - completion of training portfolios and career progression still rely on the observation of performance by relatively small numbers of colleagues. In a sense, this work alludes to the real-world dynamic between trainee and specialist educator on the hospital floor, requiring an experienced senior to recognise competence and knowledge when present in a junior, or limitations and gaps when absent.

The question of whether the specialist educator is participating in a reproducible way remains moot. Experts will differ, medicine is rarely ‘black and white’. Yet, the same is true of LLMs in which randomness is inherent; the same inputs will produce different outputs one day to the next; modeling processes evolve. Throughout, the human denominator retains influence over what is offered to the chatbot - an analysis of the effect of the initial and subsequent prompts on clinical utility is an area for future research as is the use of multiple human specialists and multiple LLMs to improve validity and encourage consensus.

To date, LLMs have been shown to successfully navigate clinical board questions and sequentially and linearly refine diagnostic possibilities when single scenario inputs are offered (1,4,5). Measuring the effectiveness of LLMs in complex and fluid clinical interactions is clearly more demanding. As outlined herein, the evaluation of ChatGPT as it learns to become a trained expert (as part of a journey towards supporting (or replacing) the specialist) is one potential method of testing and improving its clinical credibility.

## Data Availability

All data produced in the present work are contained in the manuscript

